# CARDIO-METABOLIC AND RENAL RISK AWARENESS AND SELF-MEDICATION PRACTICES AMONG COMMERCIAL BUS DRIVERS IN EBONYI STATE, NIGERIA

**DOI:** 10.64898/2026.07.26.26358985

**Authors:** Susan Chioma Udeh, Eberechukwu Blessing Nnaji, Mfreke Anakan Ayan Asigbe, Kingsley Obinna Udeh, Roland Afam Agana

## Abstract

Cardiovascular and chronic kidney disease place a growing burden on low-resource settings, and commercial drivers, whose occupation combines prolonged sitting, irregular meals and limited contact with routine health services, are an understudied high-risk group in Nigeria. Self-medication, common across the country, may compound this risk by masking symptoms or adding a nephrotoxic burden to an already strained cardio-metabolic system. This cross-sectional analytical study examined cardio-metabolic and renal risk awareness, self-medication practices and their predictors among 435 commercial bus drivers recruited from motor parks in Ebonyi State, southeast Nigeria, using a multistage sampling approach. Data were collected through a structured interviewer-administered questionnaire and direct clinical and anthropometric measurement, including blood pressure, random blood glucose, body mass index and urine protein. Self-medication was reported by 89.4% of participants, and none had health insurance coverage. Awareness of cardio-metabolic and renal risk was unevenly distributed across the sample. Current smoking independently predicted poor awareness after adjustment (adjusted odds ratio 0.46, 95% confidence interval 0.29 to 0.71), while lower educational attainment and longer driving tenure independently predicted lower odds of self-medication. Clinical measurement identified hypertension in 12.9% and diabetes range glucose in 7.1% of participants, and more than four in five of these cases had never been previously diagnosed. Obesity was present in 25.1% of drivers. These findings show that cardio-metabolic and renal disease is accumulating largely undetected in this occupational group, that self-medication is close to universal, and that health insurance coverage is absent altogether. Occupational health programmes targeting commercial drivers should combine routine screening for hypertension and diabetes with structured education on the risks of unsupervised medicine use, particularly given the high burden of undiagnosed disease revealed by direct clinical assessment in this study.

## Introduction

Cardiovascular disease remains the world’s leading cause of mortality, accounting for an estimated 19.8 million deaths in 2022, roughly one in every three deaths recorded globally that year [1,2]. The picture worsens once chronic kidney disease is added to the equation. Recent Global Burden of Disease modelling puts the number of adults living with some stage of kidney disease at 788 million worldwide as of 2023, nearly double the 1990 figure, with the condition now ranking as the ninth leading cause of death and the twelfth leading cause of disability-adjusted life years globally [3]. What ties these two organ systems together, and what makes their joint burden so difficult to manage in low-resource settings, is that they share the same upstream drivers: uncontrolled blood pressure, dysglycaemia, excess weight, physical inactivity, and tobacco and alcohol use [4,5]. A raised systolic pressure or a fasting glucose that has crept above normal rarely announces itself with symptoms early on. By the time either organ system fails visibly, the damage is frequently irreversible, which is precisely why screening and awareness, rather than treatment alone, sit at the centre of prevention strategy [6,7].

Sub-Saharan Africa carries a disproportionate share of this burden, and Nigeria sits among the countries with the highest age-standardised chronic kidney disease prevalence worldwide, alongside Iran, Haiti and a handful of others [3,8–10]. A population-based survey conducted across eight urban communities in Kwara State found an age-adjusted chronic kidney disease prevalence of 12%, with hypertension present in a quarter of adults screened and diabetes in one in twenty-five [11]. That study also noted something worth dwelling on: Nigeria, despite its population size, still has no nationally representative data on chronic kidney disease, and most of what is known comes from scattered community and hospital-based studies [11].

Commercial drivers are an understudied occupational group because the nature of their work often keeps them away from routine healthcare contact. The job keeps a person seated for hours at a stretch, exposes them to traffic stress and irregular meal timing, and often removes them from any fixed catchment area where a health worker might reasonably expect to see them again. Several African studies bear this out [12]. Among long-distance bus drivers in Lagos, hypertension was found in nearly four out of ten men screened, with diabetes present in almost one in seven, and more than half showing dyslipidaemia [13]. A later study of intra-city drivers in Ibadan reported a similar hypertension prevalence, close to thirty-six percent, in a much smaller sample [9,14]. Findings from Ghana point in the same direction; among more than five hundred inter-regional bus drivers, unrecognised hypertension was documented in almost four out of ten participants, meaning the men themselves had no idea their pressure was already elevated [15,16]. Interestingly, a South African occupational cohort recorded a comparatively lower hypertension rate of under twenty percent, and the authors attributed part of that gap to a healthy-worker selection effect, where drivers found unfit during screening simply drop out of the workforce and out of the dataset [17]. A study in Ethiopia similarly found that the prevalence of undiagnosed hypertension among long-distance bus drivers was 22.5%, associated with modifiable behavioural factors such as insufficient physical exercise, low awareness, and a high body mass index [18]. Taken together, these numbers suggest that whatever the true prevalence turns out to be locally, it is unlikely to be trivial, and the possibility of substantial underdiagnosis has to be taken seriously.

Self-medication is a widespread but often overlooked problem in Nigeria, affecting between 60% and 90% of people across different populations [19]. Self-medication with antibiotics and antimalarial drugs was found to be high among rural dwellers surveyed in Enugu State, Nigeria [20]. Evidence from studies in southwestern Nigeria indicates that self-medication is highly prevalent across various population groups [21–24]. Closer to the present study’s setting, a survey among residents of Umuahia in Abia State, conducted through a research team affiliated with a teaching hospital in Ebonyi State, documented self-medication practices during the COVID-19 period, underscoring that the behaviour is well established in the southeast geopolitical zone [25]. Why this matters for cardio-metabolic and renal risk specifically is straightforward. Several of the drug classes most commonly self-administered in Nigeria, non-steroidal anti-inflammatory drugs chief among them, are directly nephrotoxic when taken repeatedly or at high doses, and inappropriate use can also mask symptoms of an evolving cardiovascular event or interact with an existing but undiagnosed hypertensive or diabetic state. A driver who uses an over-the-counter analgesic to relieve a headache caused by undiagnosed hypertension may only be masking the symptom while the underlying condition progresses unchecked, potentially worsening damage to already vulnerable organs.

Despite this convergence of occupational exposure, cardio-metabolic vulnerability and widespread self-medication, no study to date has examined all three together among commercial drivers in Ebonyi State. Existing driver-focused research from Nigeria has concentrated on Lagos, Ibadan and the northeast, leaving the southeast, and Ebonyi specifically, without local data to guide occupational health policy or targeted screening programmes. This gap is not a minor omission. Policy responses built on data from Lagos long-distance haulage drivers may not transfer cleanly to intra-city commercial bus drivers operating under different route lengths, income structures and access to health facilities in Abakaliki and its surrounding towns. The present study was designed to close part of that gap, combining self-reported survey data with directly measured clinical and anthropometric indicators in a single sample of 435 commercial bus drivers, so that reported awareness and behaviour could be checked against objective clinical findings rather than taken at face value.

### Aim and objectives

A cross-sectional analytical study is appropriate for this investigation. A purely descriptive design would establish prevalence but would not, on its own, support the comparison of subgroups or the estimation of association between exposure and outcome. Because the study also aims to weigh the relative contribution of socio-demographic, lifestyle and clinical factors to both awareness and self-medication behaviour, and to isolate independent predictors after adjustment for confounding, an analytical extension of the cross-sectional design was necessary from the outset. The specific objectives were to:

- Assess awareness of cardio-metabolic and renal diseases among commercial bus drivers in Ebonyi State.
- Assess self-medication practices among the study population.
- Measure associated risk factors, spanning socio-demographic, lifestyle, and directly measured clinical and anthropometric variables.
- Identify predictors of awareness and self-medication practice using multivariable logistic regression.

These objectives were achieved: awareness and self-medication were both assessed and quantified, associated risk factors were measured across all three domains, and independent predictors of each outcome were identified through multivariable logistic regression, with the results and their implications set out below.

## Materials and methods

### Study design

This was a cross-sectional analytical study that used a mixed quantitative approach. Data were collected from two complementary sources: a structured interviewer-administered questionnaire on socio-demographic characteristics, lifestyle, awareness, and self-medication practices, and direct clinical and anthropometric measurements including blood pressure, random blood glucose, weight, height, waist circumference, and urine protein. Combining self-reported information with objective measurements allowed verification of participants’ reported health status against measured findings.

### Study setting

The study was conducted among commercial bus drivers operating from motor parks and garages in Ebonyi State, located in the southeast geopolitical zone of Nigeria. Abakaliki, the state capital, hosts the largest concentration of intra-state and interstate motor parks and was the primary recruitment site, supplemented by garages in other major towns within the state to reduce the risk of drawing an unrepresentative, single-location sample. The study was conducted between April and August 2024.

Study population

The target population comprised registered commercial bus drivers, male and female, actively engaged in passenger transport within or from Ebonyi State at the time of data collection.

Inclusion criteria: drivers aged 18 years and above; actively driving commercially for a minimum of six months prior to the study, to exclude very recent entrants whose occupational exposure would be minimal; and willing to provide informed consent.

Exclusion criteria: drivers who were acutely ill or otherwise unable to participate in the interview and clinical assessment on the day of data collection; and those who declined consent or withdrew before data collection was completed.

### Sample size determination

The minimum required sample size was calculated using the Cochran formula for cross-sectional studies with an infinite or unspecified population size [26]: n = Z²pq / d², where Z is the standard normal deviate at 95% confidence (1.96), p is the estimated prevalence of the outcome of interest (self-medication practice was assumed at 50% in the absence of a comparable local estimate, to maximise the calculated sample size and guard against underpowering), q is 1 minus p, and d is the acceptable margin of error, set at 0.05. This produced a minimum sample of 384 respondents. To allow for incomplete responses and questionnaires rejected at the data-cleaning stage, the estimate was inflated by approximately 15%, giving a target of roughly 450. In total, 450 questionnaires were administered; after removing entries with substantial missing clinical data, 435 were retained for final analysis, giving a response and completion rate of 96.7%. This final analytic sample of 435 is the denominator used throughout the results reported below.

### Sampling technique

A multistage sampling approach was used. In the first stage, motor parks across Ebonyi State were stratified by local government area and a subset selected purposively to capture both urban and semi-urban terminals. In the second stage, drivers present at each selected park on data-collection days were enumerated, and systematic random sampling, using a sampling interval determined by the ratio of the daily driver count to the number required from that park, was applied to select individual participants. This two-stage approach was adopted to balance representativeness against the practical difficulty of obtaining a complete sampling frame of all commercial drivers in the state, since no centralised, up-to-date register of drivers exists. Because motor parks were selected purposively rather than at random in the first stage, prevalence estimates should be interpreted as representative of drivers operating from the parks sampled rather than as a fully probability-based estimate for all commercial drivers in the state.

### Study instrument

Data were collected using a structured, interviewer-administered questionnaire developed for the study, organised into six sections. Section A captured socio-demographic characteristics (age, marital status, education, religion, monthly income, years of commercial driving, average daily driving hours, and health insurance status). Section B assessed lifestyle characteristics, including smoking, alcohol use, physical exercise frequency, dietary patterns and water intake. Section C measured cardio-metabolic and renal risk awareness across sixteen statements, each rated on a five-point Likert scale ranging from strongly disagree to strongly agree. Section D addressed self-medication practices, first establishing whether the respondent had used medicine without a prescription in the preceding six months, then, for those who answered affirmatively, probing frequency, the classes of medicine commonly used, sources of procurement, conditions self-treated, and ten further Likert-scaled statements capturing safe-use behaviours such as reading instructions, checking expiry dates and completing recommended dosages. Section E covered health history, including prior diagnoses of hypertension, diabetes and kidney disease, family history of the same conditions, recency of blood pressure, blood glucose and kidney function testing, and the participant’s self-rated overall health. Section F, completed by trained research assistants rather than self-reported, recorded clinical measurements.

### Validity and reliability of the instrument

Content validity of the instrument was established through review by experts in public health, internal medicine and pharmacy practice, and the questionnaire was piloted on a small group of drivers outside the main study area to check clarity and flow; items flagged as ambiguous during piloting were reworded before the main survey began.

Internal consistency of the two Likert-scaled subscales was assessed using Cronbach’s alpha on the full working dataset. The sixteen-item awareness subscale returned a Cronbach’s alpha of 0.733, and the ten-item self-medication practice subscale returned a Cronbach’s alpha of 0.817, both above the conventional 0.70 threshold for acceptable internal consistency [27].

### Clinical and anthropometric measurement

All clinical measurements were taken by trained research assistants using calibrated instruments, following a standardised protocol to minimise measurement error. Blood pressure was measured twice, at least five minutes apart, with the participant seated and at rest, using a validated digital or mercury sphygmomanometer with an appropriately sized cuff; both readings were recorded, and the mean of the two was taken as the participant’s blood pressure for analysis. Pulse rate was recorded at the same sitting. Weight was measured to the nearest 0.1 kg using a calibrated digital scale, with participants in light clothing and without footwear, and height was measured to the nearest 0.01 m using a stadiometer. Body mass index was derived as weight in kilograms divided by height in metres squared. Waist circumference was measured at the midpoint between the lowest rib and the iliac crest using a non-stretchable tape. Random blood glucose was determined from a capillary blood sample using a calibrated point-of-care glucometer, and the number of hours since the participant’s last meal was also recorded, since this reading is non-fasting and its interpretation depends on how recently the participant last ate; the diabetes threshold applied to this measurement should therefore be read as a screening indicator rather than a confirmed diagnosis. Urine protein was assessed by dipstick urinalysis on a spot sample and graded as negative, trace, or one to three plus; this too is a screening test that requires confirmatory quantitative testing before a diagnosis of kidney disease can be made. Participants found on assessment to have markedly elevated blood pressure, blood glucose or proteinuria were flagged for referral to the nearest health facility, in line with the referral protocol built into the questionnaire.

### Data collection procedure

Trained research assistants, familiar with both the local languages spoken in Ebonyi State and English, administered the questionnaire face to face at motor parks during periods when drivers were between trips, to minimise disruption to their work. Each interview, including clinical measurement, took approximately twenty to twenty-five minutes to complete. Data collection took place over several weeks to capture drivers across different shift patterns and to avoid oversampling from any single time of day.

### Data management

Completed questionnaires and clinical assessment forms were checked in the field by the research assistant, and where necessary the field supervisor, before the participant left the motor park, so that obvious omissions or inconsistent entries could be corrected while the driver was still available. Each participant was assigned a unique identification number at the point of data collection, and this number was used to link the self-reported questionnaire responses to the corresponding clinical and anthropometric measurements without reference to the participant’s name, consistent with the confidentiality procedure described below.

Data were entered into IBM SPSS using the coding scheme fixed at questionnaire design, so that every categorical response corresponded to a pre-defined numeric code, for example 0 for “No” and 1 for “Yes” on binary items, or ascending integer codes for ordered response options such as frequency of exercise or years of driving. Continuous clinical and anthropometric measurements were entered at the precision recorded on the assessment form: blood pressure to the nearest whole millimetre of mercury, weight to the nearest 0.1 kilogram, height to the nearest 0.01 metre, and random blood glucose to the nearest milligram per decilitre. Both readings taken for systolic and diastolic blood pressure were entered separately and retained in the dataset alongside their computed averages, so that the averaging step used in analysis remains checkable rather than taken on trust.

After entry, the dataset was screened for out-of-range and internally inconsistent values. Continuous variables were checked against plausible physiological limits, the composite awareness and practice scores were checked by re-summing their constituent Likert items and comparing the result against the recorded total, and body mass index was checked by recalculating it from the entered weight and height. Records with unresolved missing values or implausible clinical readings on key variables were excluded at this stage, which is why 15 questionnaires were dropped between the 450 administered and the 435 retained for analysis, as described under Sample size determination. The final analytic file used for this study accordingly contains no missing values on any of the variables carried into the analysis reported below.

### Measurement of variables

Variables were measured and coded at three levels in SPSS, matching the type of information each captured. Socio-demographic and lifestyle variables from Sections A and B, including age band, marital status, educational level, religion, monthly income, years of commercial driving, smoking, alcohol use, exercise frequency, fruit and vegetable intake, sugary drink consumption, adequate water intake and health insurance status, were entered as categorical codes corresponding to the fixed response options on the questionnaire. Average daily driving hours was the exception within this block, entered as a continuous count in whole hours, with an observed range of 4 to 16 hours and a mean of 10.1 hours (standard deviation 2.1), rather than as a banded category.

The sixteen awareness items in Section C and the ten safe-medication-practice items in Section D were each entered on their original five-point Likert scale, then summed to produce a continuous awareness score, with an observed range of 34 to 74 out of a possible 16 to 80, and a continuous practice score, with an observed range of 11 to 50 out of a possible 10 to 50. Each composite score was then recoded into a three-level ordinal category, low, moderate or high, using the 33rd and 66th percentile of its own distribution as the cut points; in this sample that corresponded to scores of 52 and 59 for awareness and 35 and 40 for practice. For the multivariable analysis described below, the three-level awareness category was further collapsed into a binary indicator, poor awareness, defined as the lowest tertile of the awareness score (36.1% of participants), versus moderate or high awareness combined.

Self-medication was defined as the use of any medicine without a prescription or consultation with a qualified health professional within the six months preceding the interview, and was entered as a binary variable alongside the related categorical items on frequency, the classes of medicine used, sources of procurement and conditions self-treated; this binary variable, rather than the practice score, was the outcome used in the multivariable self-medication model. The health-history items in Section E, covering prior diagnosis of hypertension, diabetes and kidney disease, whether blood pressure, blood sugar and kidney function had previously been checked, and self-rated overall health, were likewise entered as categorical codes. Family history of hypertension, diabetes and kidney disease was coded across three levels, No, Yes, or I do not know, since a meaningful share of participants were unable to answer with certainty.

Clinical and anthropometric measurements taken by the research assistants in Section F were entered as continuous variables: the two systolic and two diastolic blood pressure readings and their computed averages, pulse rate, weight, height, body mass index, waist circumference, random blood glucose and the time elapsed since the participant’s last meal. Urine protein on dipstick testing was entered as an ordinal category, negative, trace, 1+, 2+ or 3+, reflecting how the test is read rather than as a numeric concentration.

Three further clinical variables were derived directly from the measured data, independently of what participants reported about their own health, so that measured status could be compared against self-reported status rather than treated as interchangeable. Measured hypertension was defined as an average systolic blood pressure of 140 mmHg or higher, an average diastolic blood pressure of 90 mmHg or higher, or both. Obesity was defined as a body mass index of 30 kg/m² or higher. Measured diabetes was defined as a random blood glucose of 200 mg/dL or higher, consistent with conventional epidemiological cut-offs used in comparable Nigerian driver studies [8,9]. Each of these measured flags was coded as a separate variable from its self-reported counterpart, prior diagnosis of hypertension, diabetes or kidney disease, which is what allowed the study to check, for a given participant, whether a measured abnormality on the day of assessment matched what the participant believed about their own health.

### Statistical analysis

Data were entered, cleaned and analysed using IBM SPSS. Categorical variables were summarised as frequencies and percentages, while continuous variables, including average systolic and diastolic blood pressure, body mass index, random blood glucose and waist circumference, were summarised as means with standard deviations. Normality of continuous variables was assessed using the Shapiro-Wilk test together with visual inspection of distribution plots; random blood glucose showed a pronounced right skew, consistent with the non-fasting sampling protocol, and is additionally presented as a box plot alongside its mean and median for transparency.

Associations between categorical socio-demographic and lifestyle variables (age group, marital status, educational level, monthly income, years of commercial driving, smoking status, alcohol use, exercise frequency, fruit and vegetable intake, sugary drink consumption, and adequacy of water intake) and both awareness category and self-medication status were examined using the chi-square test of independence, or Fisher’s exact test where more than 20% of expected cell counts fell below five.

Pearson correlation was used to explore the linear relationship between the continuous awareness score, the continuous self-medication practice score, and the measured clinical and anthropometric parameters (random blood glucose, waist circumference, body mass index, and average systolic and diastolic blood pressure); Spearman correlation was reserved for any variable pair that departed materially from bivariate normality.

To identify independent predictors of poor cardio-metabolic and renal risk awareness and of self-medication practice, two separate binary logistic regression models were fitted, one for each outcome. Candidate covariates (age group, educational level, monthly income, years of commercial driving, fitted both categorically and as a continuous per-year term, current smoking status, and alcohol consumption) were selected using a combination of bivariate significance (p<0.20) on chi-square testing and a priori clinical relevance drawn from comparable driver-based cardio-metabolic studies, and were entered simultaneously into each model. Marital status met the bivariate significance threshold for self-medication (p=0.042) but was excluded from the final multivariable model because one category, widowed drivers (n=13), showed complete separation with respect to self-medication, as all 13 reported self-medicating, which prevented stable estimation when the four-level variable was entered alongside the other covariates. A sensitivity analysis using a collapsed married versus not-married indicator converged without difficulty and showed no independent association with self-medication after adjustment (adjusted odds ratio 0.89, 95% confidence interval 0.46 to 1.72, p=0.724), supporting the decision to exclude marital status from the reported model. Crude odds ratios from unadjusted bivariate logistic regression and adjusted odds ratios from the multivariable model, each with 95% confidence intervals, were reported for every covariate category relative to a reference group. Model fit was assessed using McFadden’s pseudo R-squared and the likelihood ratio test comparing each fitted model against the intercept-only model. The awareness model had a pseudo R-squared of 0.051 and improved significantly on the null model (likelihood ratio p=0.0065). The self-medication model had a pseudo R-squared of 0.064; although two individual predictors reached significance, the overall model did not significantly improve on the null model (likelihood ratio p=0.128), which is reported here as a limitation of the model’s overall explanatory power rather than of the individual associations identified. A p-value below 0.05 was considered statistically significant throughout, and a p-value between 0.05 and 0.10 was interpreted as a borderline or trend-level association rather than a confirmed one.

### Ethical considerations

Ethical approval for the study was obtained from the Ethics Committee of the School of Medicine, University of Nigeria Teaching Hospital (UNTH), Ituku-Ozalla, Enugu, Nigeria, with reference number UNTH/HREC/2024/03/900. The study was conducted in accordance with the principles of the Declaration of Helsinki. Permission to access motor parks was separately sought from park management and drivers’ union representatives. Participants provided written informed consent; thumb-printed consent, witnessed and documented in line with the approving committee’s requirements, was obtained from participants who could not read or write. Participation was entirely voluntary, and participants had the right to withdraw from the study at any time without penalty or any consequence for their work. Confidentiality was maintained by removing any identifying information from the questionnaires and using unique codes for each participant. Clinical findings requiring urgent attention were communicated directly to the affected participant along with a referral note to the nearest health facility.

## Results

Table 1 shows that the study population (N = 435) was predominantly young and of working age, with nearly three-quarters (71.7%) aged 18 to 37 years and only 3.4% aged 58 years or older. Slightly more than half of the respondents (52.0%) were single, while 40.0% were married. Educational attainment was generally modest, as about half (50.8%) had completed only secondary education, while just 13.8% possessed tertiary qualifications. The sample showed limited religious diversity, with Christianity accounting for 95.9% of respondents. Income levels were relatively low, with almost 60% earning less than ₦100,000 per month and only 10.8% earning ₦150,000 or more. Driving-related characteristics suggest substantial occupational road exposure, as 45.5% had less than six years of driving experience and more than half (55.4%) reported driving between 9 and 11 hours daily; average daily driving hours across the whole sample had a mean of 10.1 hours (standard deviation 2.1, range 4 to 16). A particularly notable finding was the complete absence of health insurance coverage among respondents, highlighting potential barriers to healthcare access and suggesting increased vulnerability to adverse outcomes if cardio-metabolic or renal health risks are present.

**Table 1.** Socio-demographic characteristics of respondents (N = 435).

| Variable | Category | Frequency | Percent (%) |
| --- | --- | --- | --- |
| Age | 18-27 | 127 | 29.2 |
|  | 28-37 | 185 | 42.5 |
|  | 38-47 | 70 | 16.1 |
|  | 48-57 | 38 | 8.7 |
|  | 58+ | 15 | 3.4 |
|  | Total | 435 | 100.0 |
| Marital status | Single | 226 | 52.0 |
|  | Married | 174 | 40.0 |
|  | Divorced | 22 | 5.1 |
|  | Widowed | 13 | 3.0 |
|  | Total | 435 | 100.0 |
| Highest educational qualification | No formal education | 30 | 6.9 |
|  | Primary | 124 | 28.5 |
|  | Secondary | 221 | 50.8 |
|  | Tertiary | 60 | 13.8 |
|  | Total | 435 | 100.0 |
| Religion | Christian | 417 | 95.9 |
|  | Islam | 11 | 2.5 |
|  | Traditional | 7 | 1.6 |
|  | Total | 435 | 100.0 |
| Monthly income | <₦50,000 | 67 | 15.4 |
|  | ₦50,000-99,999 | 192 | 44.1 |
|  | ₦100,000-149,999 | 129 | 29.7 |
|  | ₦150,000+ | 47 | 10.8 |
|  | Total | 435 | 100.0 |
| Years driving | Less than 6 | 198 | 45.5 |
|  | 6-8 | 116 | 26.7 |
|  | 9-11 | 83 | 19.1 |
|  | 12 or more | 38 | 8.7 |
|  | Total | 435 | 100.0 |
| Health insurance | No | 435 | 100.0 |
|  | Total | 435 | 100.0 |

Note: average daily driving hours, treated as a continuous variable, had a mean of 10.1 hours (standard deviation 2.1, range 4 to 16) and is not shown as a banded category above.

Alcohol consumption (89.9%) and self-medication (89.4%) were highly prevalent among participants, while 30.3% reported smoking. Most participants exercised rarely (39.8%) and consumed fruits and vegetables only occasionally (44.1%), although 80.7% reported adequate water intake. Sugary drink consumption was common, with 46.0% consuming them several times a week or daily. Frequent or very frequent self-medication was reported by 70.8%, with vitamins (20.0%) and malaria medicines (17.7%) being the most used medications. Previous diagnoses of hypertension, diabetes, and kidney disease were reported by 18.4%, 11.0%, and 0.9% of participants, respectively. A family history of hypertension was reported by 57.7%, while only 25.7%, 37.2%, and 9.2% had checked their blood pressure, blood sugar, and kidney function, respectively. Awareness (36.1%, 32.2%, and 31.7%) and practice (34.5%, 31.7%, and 33.8%) categories were relatively evenly distributed (Table 2).

**Table 2.** Lifestyle behaviours, self-medication practices, family history, clinical history, and awareness of cardio-metabolic and renal risk among commercial bus drivers (N = 435).

| Variable | Category | Frequency | Percent (%) |
| --- | --- | --- | --- |
| Smoking | No | 303 | 69.7 |
|  | Yes | 132 | 30.3 |
| Alcohol | No | 44 | 10.1 |
|  | Yes | 391 | 89.9 |
| Exercise | Rarely | 173 | 39.8 |
|  | Occasionally | 122 | 28.0 |
|  | Weekly | 95 | 21.8 |
|  | Daily | 45 | 10.3 |
| Fruit/vegetables | Occasionally | 192 | 44.1 |
|  | Weekly | 120 | 27.6 |
|  | Several times a week | 91 | 20.9 |
|  | Daily | 32 | 7.4 |
| Sugary drinks | Rarely | 121 | 27.8 |
|  | Weekly | 114 | 26.2 |
|  | Several times a week | 104 | 23.9 |
|  | Daily | 96 | 22.1 |
| Adequate water | No | 84 | 19.3 |
|  | Yes | 351 | 80.7 |
| Self-medication | No | 46 | 10.6 |
|  | Yes | 389 | 89.4 |
| Self-medication frequency | Rarely | 42 | 9.7 |
|  | Occasionally | 85 | 19.5 |
|  | Frequently | 172 | 39.5 |
|  | Very frequently | 136 | 31.3 |
| Common medicine | Pain relievers | 60 | 13.8 |
|  | Antibiotics | 73 | 16.8 |
|  | Malaria medicines | 77 | 17.7 |
|  | Vitamins | 87 | 20.0 |
|  | Herbal medicines | 72 | 16.6 |
|  | Others | 66 | 15.2 |
| Medicine source | Pharmacy | 87 | 20.0 |
|  | Patent medicine shop | 92 | 21.1 |
|  | Friend or family | 80 | 18.4 |
|  | Leftover medicines | 81 | 18.6 |
|  | Herbal seller | 95 | 21.8 |
| Health conditions treated | Headache | 85 | 19.5 |
|  | Fever | 49 | 11.3 |
|  | Malaria | 64 | 14.7 |
|  | Body pains | 80 | 18.4 |
|  | Stomach upset | 82 | 18.9 |
|  | Others | 75 | 17.2 |
| Hypertension diagnosis | No | 355 | 81.6 |
|  | Yes | 80 | 18.4 |
| Diabetes diagnosis | No | 387 | 89.0 |
|  | Yes | 48 | 11.0 |
| Kidney disease diagnosis | No | 431 | 99.1 |
|  | Yes | 4 | 0.9 |
| Family history hypertension | No | 98 | 22.5 |
|  | Yes | 251 | 57.7 |
|  | I do not know | 86 | 19.8 |
| Family history diabetes | No | 150 | 34.5 |
|  | Yes | 117 | 26.9 |
|  | I do not know | 168 | 38.6 |
| Family history kidney disease | No | 209 | 48.0 |
|  | Yes | 16 | 3.7 |
|  | I do not know | 210 | 48.3 |
| Blood pressure checked | No | 323 | 74.3 |
|  | Yes | 112 | 25.7 |
| Blood sugar checked | No | 273 | 62.8 |
|  | Yes | 162 | 37.2 |
| Kidney function checked | No | 282 | 64.8 |
|  | Yes | 40 | 9.2 |
|  | I do not know | 113 | 26.0 |
| Awareness category | Low | 157 | 36.1 |
|  | Moderate | 140 | 32.2 |
|  | High | 138 | 31.7 |
| Practice category | Low | 150 | 34.5 |
|  | Moderate | 138 | 31.7 |
|  | High | 147 | 33.8 |

The mean systolic blood pressure was 124.8 ± 12.7 mmHg (range: 93.0 to 162.0 mmHg), while the mean diastolic blood pressure was 73.0 ± 7.6 mmHg (range: 55.0 to 96.0 mmHg). Participants had a mean body mass index of 27.1 ± 4.5 kg/m² (range: 16.7 to 39.5 kg/m²), indicating that the average participant was overweight. The mean waist circumference was 92.0 ± 9.7 cm (range: 66.7 to 121.9 cm), suggesting a relatively high level of central adiposity in the study population (Table 3).

**Table 3.** Clinical and anthropometric characteristics of commercial bus drivers (N = 435).

| Variable | Minimum | Maximum | Mean | Standard deviation |
| --- | --- | --- | --- | --- |
| Average systolic blood pressure (mmHg) | 93.00 | 162.00 | 124.80 | 12.69 |
| Average diastolic blood pressure (mmHg) | 55.00 | 96.00 | 72.95 | 7.55 |
| Body mass index (kg/m<sup>2</sup>) | 16.70 | 39.50 | 27.12 | 4.51 |
| Waist circumference (cm) | 66.70 | 121.90 | 91.97 | 9.72 |

##Fig 1 shows that most participants had negative urine protein dipstick results (80.0%), indicating no detectable proteinuria. A smaller proportion had trace proteinuria (12.2%), while 6.2% had 1+ protein, 0.9% had 2+ protein, and 0.7% had 3+ protein. The histogram demonstrates a markedly right-skewed distribution, with most observations concentrated at the negative category and progressively fewer participants exhibiting increasing levels of proteinuria. Overall, 20.0% of participants had detectable urinary protein (trace or higher), suggesting that a notable minority may have early or established renal abnormalities requiring further clinical evaluation.

**Fig 1.**
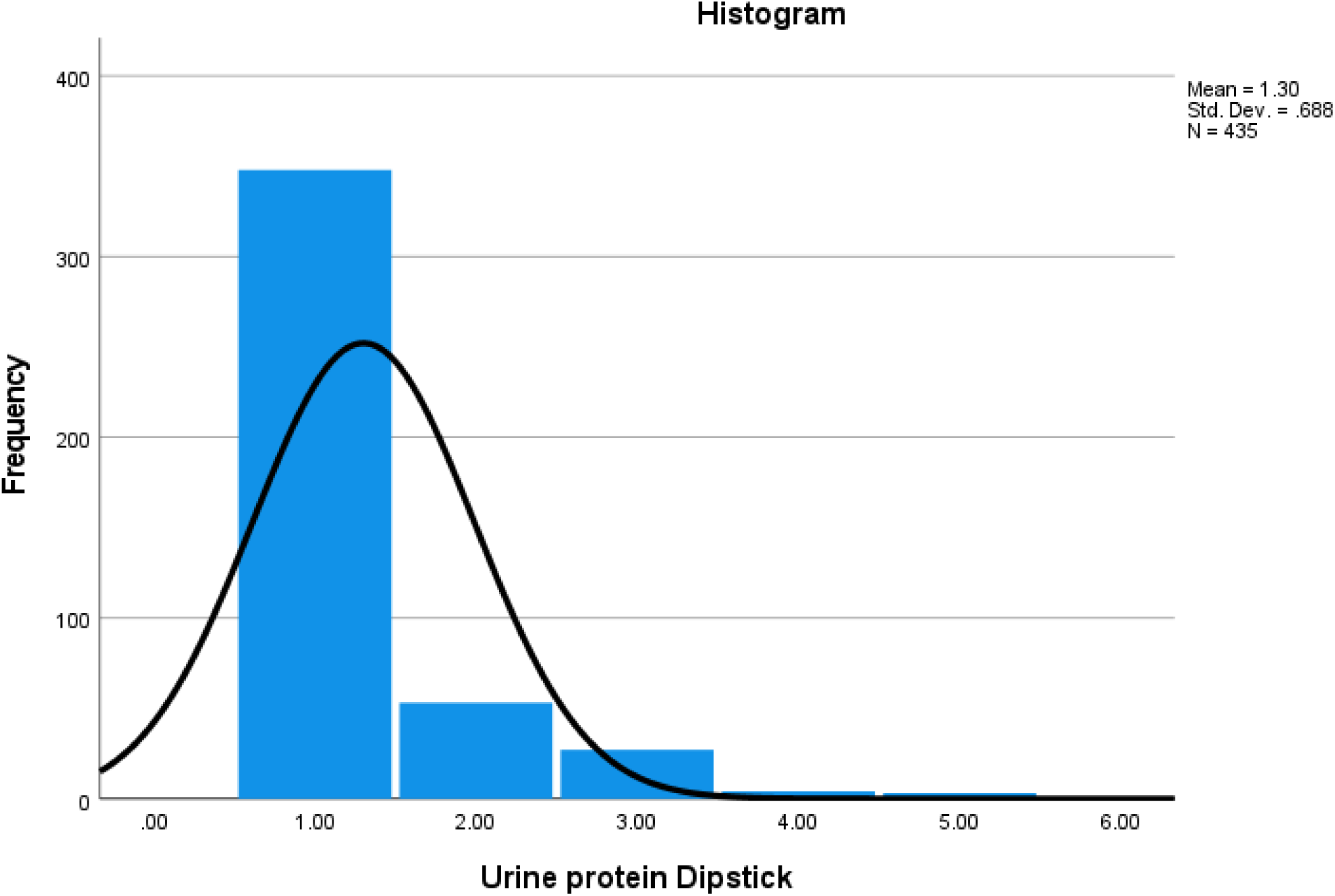
**Distribution of urine protein dipstick results among commercial bus drivers.**

According to Fig 2, random blood glucose shows a clear right skew. The mean (111.5 mg/dL) sits above the median (106 mg/dL), and the upper whisker stretches much further than the lower one, reaching as high as 241 mg/dL. This long upper tail suggests a subset of participants with notably elevated readings pulling the average up, which is common for non-fasting glucose since a recent meal or snack can push values well above what would be seen after fasting.

**Fig 2.**
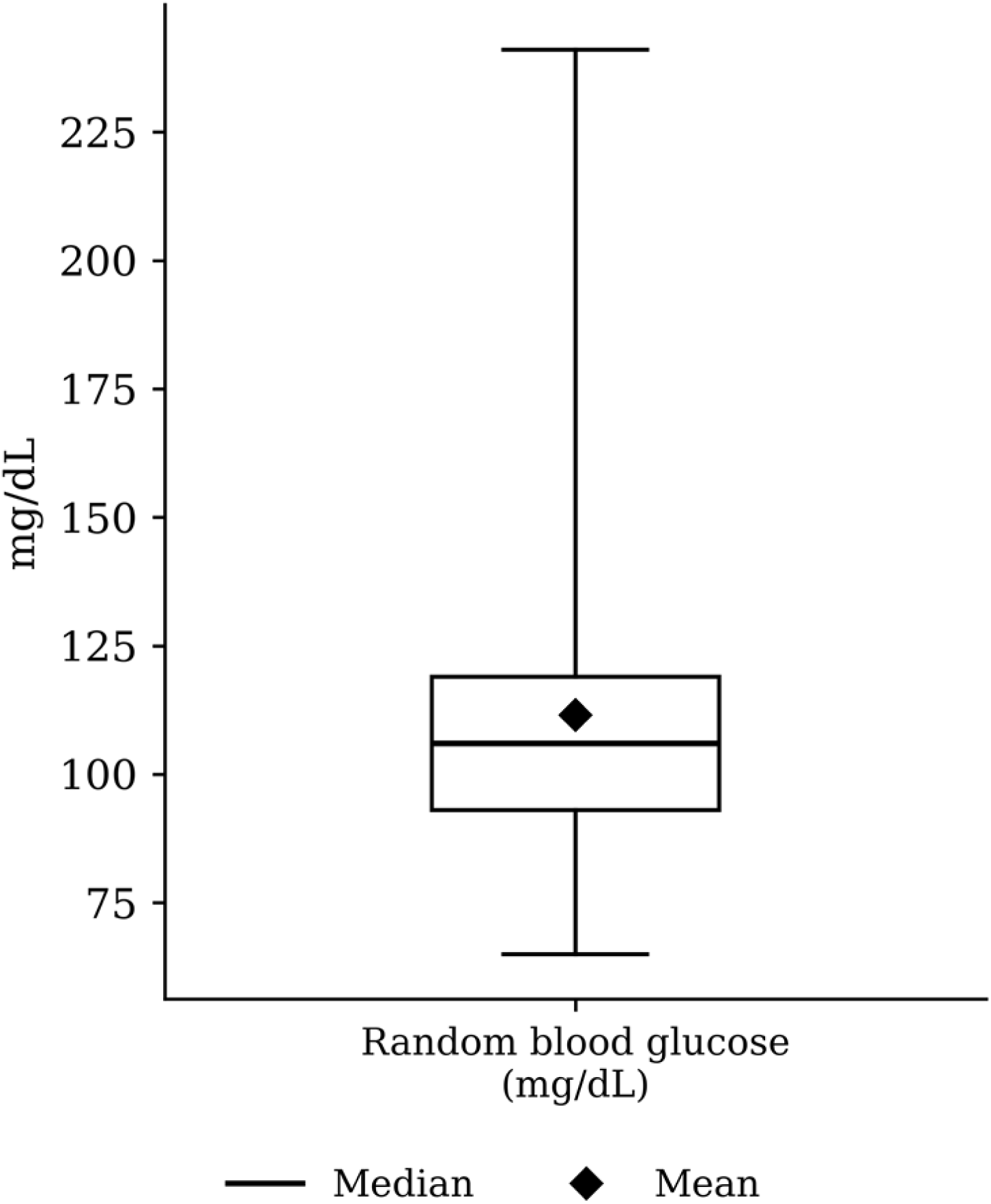
**Box plot of random blood glucose (mg/dL). The box spans the interquartile range, the horizontal line marks the median, whiskers extend to the minimum and maximum, and the diamond marks the mean.**

### Measured versus self-reported cardio-metabolic and renal disease status

Comparing measured clinical status against self-reported diagnosis revealed a substantial gap between the two. Measured hypertension (an average systolic blood pressure of at least 140 mmHg, an average diastolic blood pressure of at least 90 mmHg, or both) was present in 12.9% of participants (56 of 435), of whom 80.4% (45 of 56) had never previously been diagnosed with hypertension. Measured diabetes, defined as a random blood glucose of 200 mg/dL or higher, was present in 7.1% of participants (31 of 435), of whom 80.6% (25 of 31) had never previously been diagnosed with diabetes. Obesity, defined as a body mass index of 30 kg/m² or higher, was present in 25.1% of participants (109 of 435). These figures indicate that most of the cardio-metabolic disease present in this workforce on the day of assessment had not previously come to clinical attention (Table 4).

**Table 4.** Measured versus self-reported cardio-metabolic and renal disease status (N = 435).

| Measured condition | Prevalence, n (%) | Previously diagnosed, n (%) | Undiagnosed, n (%) |
| --- | --- | --- | --- |
| Hypertension (SBP ≥140 mmHg or DBP ≥90 mmHg) | 56 (12.9) | 11 (19.6) | 45 (80.4) |
| Diabetes (random glucose ≥200 mg/dL) | 31 (7.1) | 6 (19.4) | 25 (80.6) |
| Obesity (BMI ≥30 kg/m<sup>2</sup>) | 109 (25.1) | N/A | N/A |

*Note: previously diagnosed and undiagnosed percentages are expressed as a share of participants meeting the measured threshold for that condition. Obesity has no directly comparable self-reported diagnosis item in the questionnaire, so previously diagnosed and undiagnosed shares are not applicable (N/A). SBP, systolic blood pressure; DBP, diastolic blood pressure; BMI, body mass index.*

According to Table 5, only three associations reached statistical significance (p<0.05): smoking status was strongly associated with awareness category (χ² = 16.09, p < 0.001), while marital status (χ² = 8.18, p = 0.042) and years of driving experience (χ² = 11.41, p = 0.010) were significantly associated with self-medication behaviour; education showed a borderline, non-significant trend with self-medication (p = 0.098). All other demographic and lifestyle variables tested, age, income, alcohol use, exercise, diet (fruit and vegetables, sugary drinks), water intake, religion, and health insurance, showed no statistically significant relationship with either awareness category or self-medication, indicating that in this sample of 435 respondents, awareness of cardio-metabolic and renal risk and self-medication practices were largely independent of most demographic and behavioural characteristics, with smoking, marital status, and driving experience being notable exceptions. As described in the statistical analysis, the association with marital status was driven in large part by a small, fully self-medicating widowed subgroup and did not persist in a collapsed, model-stable form.

**Table 5.** Associations between demographic and lifestyle variables and health awareness category and self-medication practice (N = 435).

| Variable | Awareness $\chi^2$ | Awareness p-value | Self-medication $\chi^2$ | Self-medication p-value |
| --- | --- | --- | --- | --- |
| Age | 8.080 | 0.426 | 1.527 | 0.822 |
| Education | 4.999 | 0.544 | 6.303 | 0.098 |
| Marital status | 3.551 | 0.737 | 8.181 | 0.042 |
| Monthly income | 6.134 | 0.408 | 0.970 | 0.808 |
| Years driving | 7.897 | 0.246 | 11.412 | 0.010 |
| Smoking | 16.091 | <0.001 | 0.479 | 0.489 |
| Alcohol use | 0.003 | 0.998 | 0.114 | 0.736 |
| Exercise frequency | 5.068 | 0.535 | 2.690 | 0.442 |
| Fruit/vegetable intake | 5.861 | 0.439 | 6.046 | 0.109 |
| Sugary drinks | 4.505 | 0.609 | 4.128 | 0.248 |
| Adequate water intake | 0.039 | 0.981 | 0.553 | 0.457 |

Table 6 showed a moderate positive relationship between awareness and preventive practice scores (r = 0.487, p < 0.001), indicating that participants with greater awareness were more likely to engage in better preventive practices. Awareness was weakly associated with waist circumference (r = 0.100, p = 0.037) but showed no significant correlations with blood glucose, body mass index, or blood pressure. Preventive practice scores were not significantly correlated with any clinical risk indicator. Random blood glucose was weakly but significantly associated with waist circumference (r = 0.175, p < 0.001) and body mass index (r = 0.191, p < 0.001). Waist circumference demonstrated a very strong positive correlation with body mass index (r = 0.918, p < 0.001) and moderate positive correlations with systolic (r = 0.510, p < 0.001) and diastolic blood pressure (r = 0.425, p < 0.001). Similarly, body mass index was moderately correlated with systolic (r = 0.532, p < 0.001) and diastolic blood pressure (r = 0.448, p < 0.001). Systolic and diastolic blood pressure were weakly but significantly correlated (r = 0.163, p = 0.001). These findings indicate that anthropometric measures are strongly related to blood pressure and glucose levels, whereas awareness and preventive practices have limited direct associations with the measured cardio-metabolic risk indicators.

**Table 6.** Pearson correlation matrix showing relationships between awareness, preventive practice, and cardio-metabolic risk indicators (N = 435).

| Variables | Awareness | Practice | Glucose | Waist | BMI | SBP | DBP |
| --- | --- | --- | --- | --- | --- | --- | --- |
| Awareness | 1 | 0.487 (<0.001) | 0.012 (0.806) | 0.100 (0.037) | 0.068 (0.154) | 0.010 (0.833) | 0.048 (0.318) |
| Practice | 0.487 (<0.001) | 1 | 0.011 (0.824) | 0.004 (0.936) | -0.018 (0.704) | -0.063 (0.188) | -0.064 (0.184) |
| Glucose | 0.012 (0.806) | 0.011 (0.824) | 1 | 0.175 (<0.001) | 0.191 (<0.001) | 0.041 (0.392) | 0.060 (0.211) |
| Waist | 0.100 (0.037) | 0.004 (0.936) | 0.175 (<0.001) | 1 | 0.918 (<0.001) | 0.510 (<0.001) | 0.425 (<0.001) |
| BMI | 0.068 (0.154) | -0.018 (0.704) | 0.191 (<0.001) | 0.918 (<0.001) | 1 | 0.532 (<0.001) | 0.448 (<0.001) |
| SBP | 0.010 (0.833) | -0.063 (0.188) | 0.041 (0.392) | 0.510 (<0.001) | 0.532 (<0.001) | 1 | 0.163 (0.001) |
| DBP | 0.048 (0.318) | -0.064 (0.184) | 0.060 (0.211) | 0.425 (<0.001) | 0.448 (<0.001) | 0.163 (0.001) | 1 |

Note: values are Pearson correlation coefficients, with p-values in parentheses. BMI, body mass index; SBP, average systolic blood pressure; DBP, average diastolic blood pressure.

Table 7 shows the factors associated with poor awareness using crude and adjusted logistic regression analyses. After adjustment, current smoking remained the only significant predictor, with non-smokers having significantly lower odds of poor awareness compared with smokers (adjusted odds ratio 0.455, 95% confidence interval 0.293 to 0.708, p < 0.001). Age, educational level, monthly income, years of driving, and alcohol consumption were not significantly associated with poor awareness after controlling for other variables. Although years of driving showed a borderline association, with each additional year increasing the odds of poor awareness by approximately 23% (adjusted odds ratio 1.226, 95% confidence interval 0.999 to 1.506, p = 0.052), this did not reach statistical significance. Overall, smoking status was the main independent factor associated with poor awareness in this model, which showed an acceptable overall fit (McFadden’s pseudo R-squared 0.051, likelihood ratio p = 0.0065).

**Table 7.** Predictors of poor awareness among study participants (N = 435).

| Variable | Category | COR | 95% CI | p | AOR | 95% CI | p |
| --- | --- | --- | --- | --- | --- | --- | --- |
| Age | 18-27 | 0.388 | 0.131-1.144 | 0.086 | 0.464 | 0.149-1.442 | 0.184 |
|  | 28-37 | 0.583 | 0.203-1.677 | 0.317 | 0.707 | 0.232-2.154 | 0.542 |
|  | 38-47 | 0.457 | 0.148-1.411 | 0.173 | 0.537 | 0.164-1.761 | 0.305 |
|  | 48-57 | 0.404 | 0.119-1.373 | 0.146 | 0.532 | 0.147-1.929 | 0.337 |
|  | 58+ (ref) | 1 | - | - | 1 | - | - |
| Education | No formal education | 0.727 | 0.275-1.920 | 0.520 | 0.650 | 0.236-1.787 | 0.404 |
|  | Primary | 1.493 | 0.784-2.843 | 0.223 | 1.455 | 0.741-2.856 | 0.276 |
|  | Secondary | 1.048 | 0.573-1.918 | 0.878 | 1.036 | 0.549-1.956 | 0.912 |
|  | Tertiary (ref) | 1 | - | - | 1 | - | - |
| Monthly income | <₦50,000 | 1.035 | 0.451-2.377 | 0.935 | 1.046 | 0.442-2.473 | 0.919 |
|  | ₦50,000-99,999 | 1.828 | 0.907-3.685 | 0.091 | 1.581 | 0.762-3.280 | 0.219 |
|  | ₦100,000-149,999 | 1.449 | 0.696-3.019 | 0.321 | 1.408 | 0.655-3.027 | 0.381 |
|  | ₦150,000+ (ref) | 1 | - | - | 1 | - | - |
| Years driving (per year) | Continuous | 1.174 | 0.966-1.427 | 0.107 | 1.226 | 0.999-1.506 | 0.052 |
| Currently smokes | No | 0.449 | 0.295-0.684 | <0.001 | 0.455 | 0.293-0.708 | <0.001 |
|  | Yes (ref) | 1 | - | - | 1 | - | - |
| Drinks alcohol | No | 1.013 | 0.530-1.937 | 0.968 | 1.017 | 0.511-2.024 | 0.962 |
|  | Yes (ref) | 1 | - | - | 1 | - | - |

Note: COR, crude odds ratio; AOR, adjusted odds ratio; CI, confidence interval; ref, reference category. Model fit: McFadden's pseudo R-squared = 0.051; likelihood ratio test vs null model, $p = 0.0065$ .

Table 8 shows the factors associated with self-medication practice using crude and adjusted logistic regression analyses. After adjustment, education level and years of driving were significantly associated with self-medication. Participants with no formal education (adjusted odds ratio 0.094, 95% confidence interval 0.010 to 0.926, p = 0.043) and those with secondary education (adjusted odds ratio 0.112, 95% confidence interval 0.015 to 0.856, p = 0.035) had significantly lower odds of self-medication compared with those with tertiary education. Additionally, each additional year of driving was associated with a reduction in the odds of self-medication (adjusted odds ratio 0.681, 95% confidence interval 0.505 to 0.918, p = 0.012). Age, monthly income, smoking status, and alcohol consumption were not significantly associated with self-medication after controlling for other variables. Overall, education level and duration of driving were the main independent factors influencing self-medication practices in this study. The model as a whole had a modest fit (McFadden’s pseudo R-squared 0.064), and the overall likelihood ratio test did not reach significance (p = 0.128) despite the two individual predictors described above; this should be borne in mind when interpreting the strength of the model as a whole, as distinct from the two significant individual associations it identified.

**Table 8.** Predictors of self-medication practice among study participants (N = 435).

| Variable | Category | COR | 95% CI | p | AOR | 95% CI | p |
| --- | --- | --- | --- | --- | --- | --- | --- |
| Age | 18-27 | 1.242 | 0.254-6.083 | 0.789 | 1.054 | 0.208-5.345 | 0.949 |
|  | 28-37 | 1.625 | 0.337-7.846 | 0.546 | 1.438 | 0.285-7.249 | 0.660 |
|  | 38-47 | 1.043 | 0.201-5.402 | 0.960 | 0.792 | 0.145-4.331 | 0.788 |
|  | 48-57 | 1.015 | 0.175-5.907 | 0.986 | 0.995 | 0.160-6.188 | 0.996 |
|  | 58+ (ref) | 1 | - | - | 1 | - | - |
| Education | No formal education | 0.110 | 0.012-1.034 | 0.054 | 0.094 | 0.010-0.926 | 0.043 |
|  | Primary | 0.145 | 0.018-1.134 | 0.066 | 0.157 | 0.020-1.248 | 0.080 |
|  | Secondary | 0.117 | 0.016-0.877 | 0.037 | 0.112 | 0.015-0.856 | 0.035 |
|  | Tertiary (ref) | 1 | - | - | 1 | - | - |
| Monthly income | <₦50,000 | 1.254 | 0.393-4.003 | 0.702 | 1.444 | 0.432-4.829 | 0.550 |
|  | ₦50,000-99,999 | 1.131 | 0.431-2.968 | 0.803 | 1.295 | 0.472-3.554 | 0.616 |
|  | ₦100,000-149,999 | 1.570 | 0.546-4.514 | 0.403 | 1.708 | 0.569-5.130 | 0.340 |
|  | ₦150,000+ (ref) | 1 | - | - | 1 | - | - |
| Years driving (per year) | Continuous | 0.677 | 0.507-0.904 | 0.008 | 0.681 | 0.505-0.918 | 0.012 |
| Currently smokes | No | 1.255 | 0.659-2.391 | 0.489 | 1.515 | 0.761-3.017 | 0.237 |
|  | Yes (ref) | 1 | - | - | 1 | - | - |
| Drinks alcohol | No | 1.203 | 0.410-3.531 | 0.736 | 1.102 | 0.353-3.445 | 0.867 |
|  | Yes (ref) | 1 | - | - | 1 | - | - |

Note: COR, crude odds ratio; AOR, adjusted odds ratio; CI, confidence interval; ref, reference category. Marital status was assessed but excluded from this model; see Materials and methods for details. Model fit: McFadden's pseudo R-squared = 0.064; likelihood ratio test vs null model, $p = 0.128$ .

## Discussion

This study set out to characterise cardio-metabolic and renal risk awareness and self-medication behaviour among 435 commercial bus drivers in Ebonyi State, and to identify the socio-demographic, lifestyle, and clinical correlates of both. The single most striking result to emerge is the near-universal recourse to unsupervised medicine use: almost nine in ten drivers reported self-medicating in the six months preceding interview, and among those who did so, seven in ten described the practice as frequent or very frequent. Read alongside a complete absence of health insurance coverage across the entire sample, this pattern cannot be reduced to a habit picked up individually by drivers; it reads instead as the predictable outcome of an occupational group left, structurally, with self-medication as its default channel of contact with pharmacotherapy. The distribution of medicine sources reported by participants reinforces this reading: only one in five obtained medicine from a pharmacy, while patent medicine shops, herbal sellers, and friends or family together accounted for the majority of supply, a pattern consistent with an informal drug economy operating at the motor park itself, filling a gap left by the absence of any occupational health service reaching this workforce.

This finding sits alongside a comparably high figure recently reported among adults attending a teaching hospital in Ogbomosho, southwest Nigeria [21], and above the substantial share of long-term self-medicators documented in Karu, Nasarawa State [28], reinforcing that limited healthcare access, consultation cost, and unregulated over-the-counter availability, rather than any single population’s ignorance, drive this behaviour nationally. It is considerably higher than the prevalence recorded during a COVID-specific recall window in neighbouring Umuahia [25], a gap best attributed to that study’s narrower disease-specific definition rather than to any true behavioural divergence. Clinically, this matters because the conditions most commonly self-treated here, headache, body pains, and stomach upset, are precisely those for which analgesics and non-steroidal anti-inflammatory drugs are reached for first, and these agents are directly nephrotoxic with repeated or high-dose use, acting through prostaglandin inhibition, renal vasoconstriction, and, over time, interstitial nephritis and glomerular filtration decline [29]. In a sample already carrying an overweight average body mass index and a mean systolic pressure approaching the hypertensive threshold, unsupervised analgesic use functions as a second insult layered onto an already strained cardio-metabolic and renal system.

Awareness was less uniformly poor than self-medication was uniformly high, but a substantial minority of drivers still fell into the lowest awareness tertile, and only current smoking independently predicted poor awareness after adjustment. This apparent gap looks considerably worse when set against the much higher hypertension and diabetes awareness reported among adults in Imo and Kaduna states [30], but that study used a simple binary recognition question, whereas the present composite scale probes working knowledge of risk factors and complications, a far more demanding construct. This measurement gap is corroborated by a South African study that found only moderate hypertension knowledge alongside markedly better attitudes and merely fair preventive practice, explicitly naming this disconnect a knowledge-practice gap [31], a pattern this study’s own data reproduce almost exactly: awareness and safe-practice scores correlated only moderately with each other and barely at all with directly measured clinical status, while the clinical and anthropometric variables correlated strongly among themselves. These anthropometric-blood pressure correlations are considerably stronger than those reported between body mass index and blood pressure in a large Lagos community sample [32], plausibly because the present cohort is a single, occupationally homogeneous group sharing the same sedentary, long-hour driving exposure that a broader community sample dilutes.

Two predictors of self-medication were genuinely counter-intuitive. Drivers with no formal or secondary education had markedly lower odds of self-medicating than tertiary-educated peers, and each additional year of driving experience further reduced the odds. The education finding mirrors a knowledge-attitude paradox recently documented among Ecuadorian university students, where those with the greatest pharmacological knowledge self-medicated most, driven by an overconfidence effect rather than by ignorance [33]; an equally plausible alternative, given that herbal sellers and herbal remedies accounted for a substantial share of medicine sources in this sample, is that less-educated drivers are self-treating through channels the instrument did not fully capture rather than behaving more safely. The protective effect of tenure echoes the healthy-worker explanation proposed for a comparatively low undiagnosed-hypertension rate among South African bus drivers, where less healthy or less cautious drivers were thought to leave the workforce over time [17]; this selection logic plausibly extends to self-medication behaviour as well. Notably, education and tenure otherwise behave in opposite directions across the driver literature: a recent Ibadan transport-worker study found a substantial hypertension burden overall, with lower educational attainment linked to higher hypertension risk [34], the reverse of the pattern found here for self-medication, underscoring that the same socio-demographic variable can point in opposite directions depending on which cardio-metabolic outcome is examined.

Finally, the complete absence of health insurance, low screening uptake across blood pressure, glucose, and kidney function testing, and a non-trivial prevalence of detectable proteinuria, set against very low rates of prior diagnosed hypertension, diabetes, and kidney disease, together indicate substantial undetected disease. Nigeria’s National Health Insurance Scheme nominally covers outpatient blood pressure and glucose testing, but this coverage functions almost entirely within the formal economy, structurally excluding informal-sector workers such as commercial drivers, which is precisely the gap this sample’s universal lack of coverage reflects. The proteinuria finding, while requiring confirmatory testing before being read as a chronic kidney disease estimate, is consistent with the broader occupational pattern: a study using serum creatinine and urine albumin-creatinine ratio among Lagos long-haul drivers found a markedly higher chronic kidney disease burden [13] than that reported in a general Kwara State population survey using a comparable community design [11], and considerably higher than the present study’s cruder dipstick estimate, a difference that likely reflects both the more sensitive biomarker panel used in Lagos and the greater cumulative occupational exposure of long-haul relative to intra-state driving. Central adiposity and hypertension clustering documented among South African truck drivers [35] further supports reading these renal and cardiovascular signals as part of a single occupational risk syndrome rather than as isolated findings.

The direct comparison between measured and self-reported disease status in this study adds a further, more concrete dimension to this picture. More than four in five drivers found to have hypertension or diabetes-range glucose on the day of assessment had never previously been told they had the condition, and one in four drivers met the threshold for obesity. Given that health insurance coverage was entirely absent in this sample, these figures suggest that detection of cardio-metabolic disease in this occupational group currently depends almost entirely on chance contact with a health facility rather than on any structured screening pathway, a pattern consistent with the broader gap between awareness and measured status already described above. This is, to our knowledge, the first study among Nigerian commercial drivers to quantify that gap directly through paired measurement and self-report in the same sample.

### Limitations

This study has several limitations, the use of single screening measurements for diabetes and proteinuria rather than confirmed diagnoses, potential recall and social desirability bias in self-reported data, and the inability of the cross-sectional design to establish causal relationships. Additionally, the self-medication model explained only a small proportion of the observed behaviour, suggesting that other unmeasured factors may play a more important role.

## Conclusion

Taken together, these findings depict a workforce in which awareness of cardio-metabolic and renal risk is unevenly distributed and only loosely coupled to objectively measured status, in which self-medication is close to universal and shaped more by occupational structure and tenure than by ignorance, and in which a complete absence of health insurance leaves disease to be detected, if at all, by chance. The high proportion of undiagnosed hypertension and diabetes identified through direct clinical measurement in this study underscores that awareness-raising alone will not be sufficient without an accompanying structured screening programme. By extending the driver-focused cardio-metabolic literature, previously concentrated in Lagos, Ibadan, Ghana and South Africa, to Nigeria’s southeast, and by linking this for the first time to directly measured renal and clinical status, this study shows that awareness-raising and unsupervised medicine use must be addressed together, backed by direct screening, if the risk quietly accumulating among Ebonyi’s commercial drivers is to be caught before it becomes irreversible.

## Data Availability

The datasets used and/or analysed during the current study are available from the corresponding author on reasonable request.

## Acknowledgments

The authors thank the commercial bus drivers who participated in this study for their time and cooperation, and the motor park management and drivers’ union representatives in Ebonyi State who granted access to the parks and facilitated recruitment.

## Author contributions

Conceptualization: Susan Chioma Udeh. Formal analysis: Susan Chioma Udeh.

Investigation: Eberechukwu Blessing Nnaji, Kingsley Obinna Udeh.

Methodology: Mfreke Anakan Ayan Asigbe, Roland Afam Agana.

Supervision: Roland Afam Agana.

Writing, original draft: Susan Chioma Udeh, Eberechukwu Blessing Nnaji.

Writing, review and editing: Susan Chioma Udeh, Eberechukwu Blessing Nnaji, Mfreke Anakan

Ayan Asigbe, Kingsley Obinna Udeh, Roland Afam Agana.

